# Personal space Increases during the COVID-19 Pandemic in Response to Real and Virtual Humans

**DOI:** 10.1101/2021.06.09.21258234

**Authors:** Daphne J. Holt, Sarah Zapetis, Baktash Babadi, Roger B. H. Tootell

## Abstract

Typically, people maintain a certain distance from others (“personal space”) during daily life, in a largely automatic, unconscious manner. However during the COVID-19 pandemic, social distancing recommendations led to deliberate expansions of personal space outside of intimate social circles. In the laboratory, personal space preferences are quite stable over repeated measurements. Here, we collected such measurements both before and during the pandemic in the same individuals, using both conventional and virtual reality-based techniques. We found that the size of personal space, and discomfort ratings in response to personal space intrusions, increased significantly during the COVID-19 pandemic, in response to both real humans and virtual “others”. Moreover, this increase in personal space requirements correlated with the perceived, not the actual, risk of being infected with COVID-19 – even in a virtual reality environment in which there was no possibility of infection. Thus, quantification of personal space may reveal some of the psychological effects of the pandemic, and subsequent progress towards recovery.

## Introduction

“Personal space” has been defined as the area surrounding the body “into which others cannot intrude without arousing discomfort” (1). The size of personal space is moderately influenced by a number of situational and social factors, including cultural differences and gender (2-6). Despite these influences, a given individual’s preferred personal space size remains generally stable over repeated measurements under controlled circumstances (7, 8). However, such reliable personal space preferences within a given individual vary substantially *across* individuals, typically between 50 and 120 cm (4, 8, 9). Expectations about typical personal space dimensions affect many man-made aspects of our daily life, such as the architectural design of public buildings, elevator dimensions, the spacing of interior seating and queues, etc.

However during the past year, these largely unconscious personal space boundaries have been affected by “social distancing” recommendations that aimed to reduce transmission of the COVID-19 virus. These consciously adopted distances (usually 6 feet in the US, and 2 meters elsewhere) are much larger than those generated by the intrinsic brain mechanisms involved in personal space regulation. However, it is unknown whether the ongoing practice of social distancing has influenced personal space in a pervasive manner, even in virus-free contexts. To examine this question, we measured personal space size, as well as discomfort elicited by personal space intrusions, in the same individuals at two time points: *before* and *during* the COVID-19 pandemic.

## Materials and Methods

We used a well-validated experimental procedure for measuring personal space size, the Stop Distance Procedure (SDP) (9, 10). The SDP measures the distance at which subjects first become uncomfortable (the personal space boundary) when another person approaches them (passive trials), or when the subject approaches another person (active trials).

To control additional variables that could potentially influence personal space size, we also collected personal space measurements at both time points using an immersive virtual reality (VR) version of the SDP. This VR procedure measured personal space to virtual images of humans (“avatars”) but was otherwise identical to the SDP conducted with real humans. VR-based measurements of personal space can correspond quite closely to those measured to real humans, *in vivo* (3, 6, 8). Responses to personal space intrusions (discomfort ratings) were also measured at different distances within (as well as outside of) the personal space boundary, to both real and virtual humans (Figure 1).

**Figure 1.**
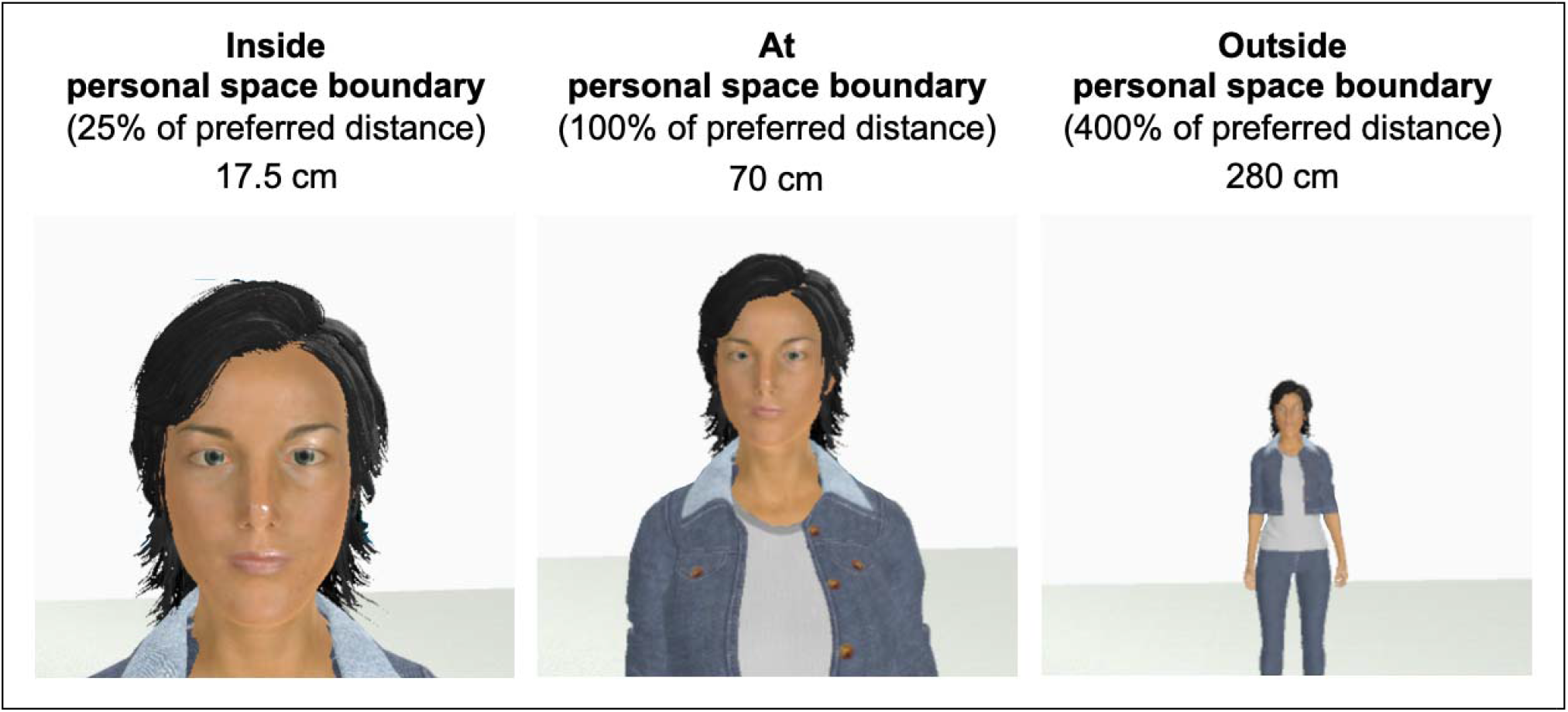
An example of an avatar used in the personal space measurements.

These measurements were first collected before the COVID-19 pandemic (September 2019 -early March 2020) in 19 subjects (47 % female, mean age = 30.6 ± 11.3 years). These measurements were then repeated in a subset of this group (n=12) after March 13th, 2020, when social distancing measures and other restrictions related to the COVID-19 pandemic were instituted in Boston, MA, where this study was conducted. During the second session, we also measured beliefs and experiences related to the COVID-19 pandemic (11). Written informed consent was obtained from all subjects prior to enrollment, in accordance with the guidelines of the Partners Healthcare Institutional Review Board which approved this study. See Supplementary Materials for additional methodological details.

## Results

As expected, the size of personal space in response to real humans was highly correlated with the size of personal space measured to virtual humans (avatars) across individuals, in both passive and active trials, both before the pandemic (passive trials: *r*(10)= .724, *p*< .001; active trials: *r*(10)= .697, *p*= .001) and during the pandemic (passive trials: *r*(10)= .928, *p*< .001; active trials: *r*(10)= .849, *p*< .001).

We found that the size of personal space was significantly larger during the COVID-19 pandemic, in response to both real and virtual humans, compared to before the pandemic (Real: passive trials: *t*(11)= 5.732, *p* < .001; active trials: *t*(11)= 3.863, *p*=.003; Virtual: passive trials: *t*(11)= 2.918, *p*= .014; active trials: *t*(11)= 3.082, *p*= .01; see Fig. 2A). In contrast, control measurements collected at two timepoints spanning a similar length of time in a prior sample (i.e. both before the COVID-19 pandemic), showed no such increases in personal space (*t*(8) = .478, *p*= .646).

**Figure 2.**
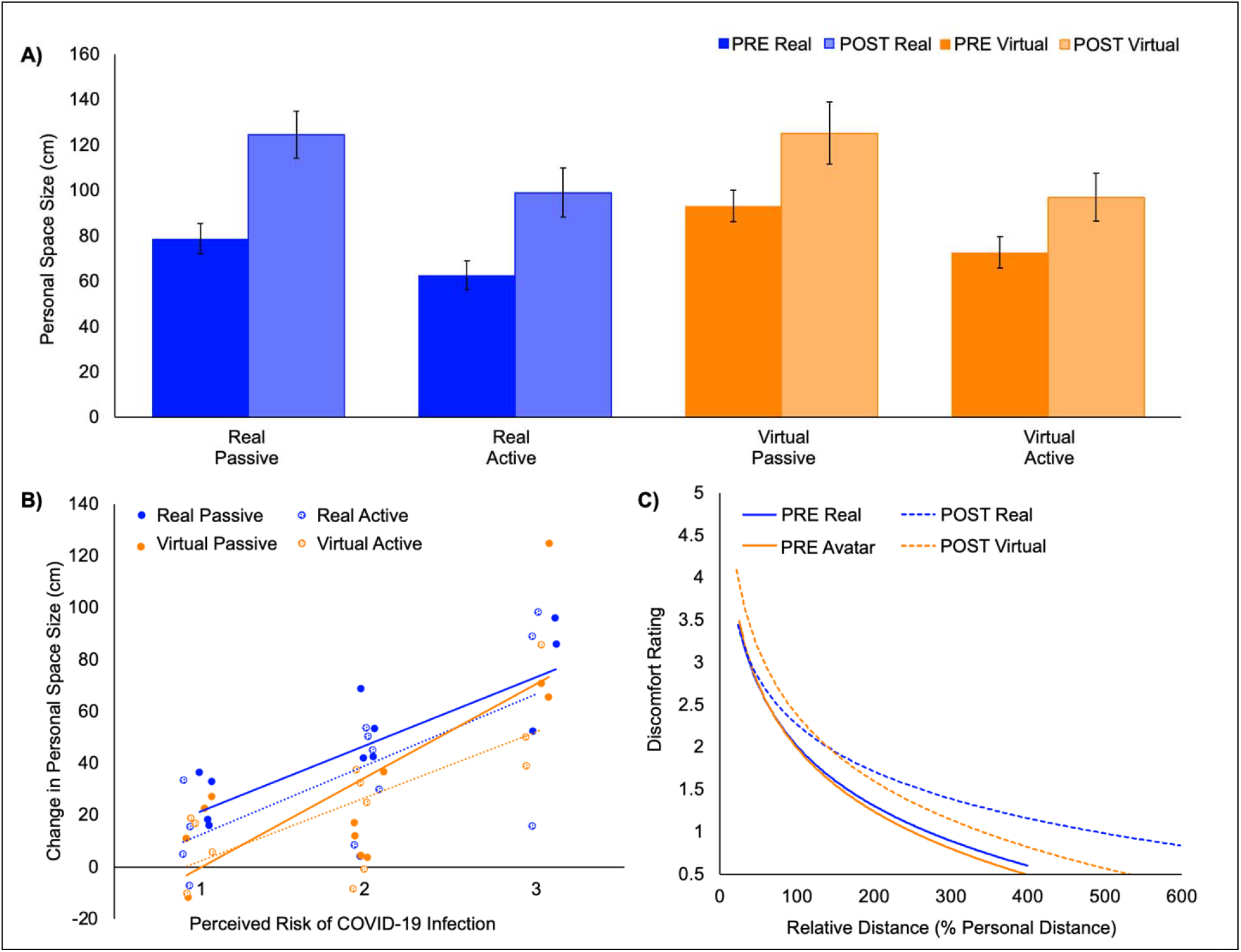
Personal space size, and discomfort during personal space intrusions, increased during the COVID-19 pandemic. **A)** Personal space size, as measured using the Stop Distance Procedure (SDP) to real and virtual humans, increased significantly during the COVID-19 pandemic. The increases in personal space during the pandemic to the real and virtual humans were highly correlated with each other (all *r* > .780; all *p* < .003). PRE = measurements collected before the COVID-19 pandemic; POST = measurements collected following the beginning of the COVID-19 pandemic. **B)** Across all four personal space measurements (i.e., real and virtual, passive and active SDP trials), the change in personal space size that occurred following the onset of the pandemic (POST -PRE) was significantly positively correlated with perceived risk of COVID-19 infection (Real: passive: *r*(10)= .732, *p*= .007, active: *r*(10)= .676, *p*= .016; Virtual: passive: *r*(10)= .736, *p*= .006, active: *r*(10)= .703, *p*= .001). **C)** Power law fits to the pre- and post-pandemic discomfort ratings, as a function of distance from real or virtual humans, expressed as percentages of pre-pandemic personal space size.

In addition, we found that the increase in personal space size during the pandemic in response to both real and virtual humans was significantly correlated with the *perceived* risk of being infected with the COVID-19 virus (ratings of “How likely do you think it is that you might become infected with COVID-19 in the near future?”; all *r* > .676; all *p* < .016; Fig. 2B). In contrast, there were no correlations between the increase in personal space size during the pandemic and levels of *actual* infection risk for each subject (as indicated by the positive COVID-19 case rate during the previous two weeks in the town in which the subject lived, all *p* > .337). Also, ratings of pandemic-related anxiety and distress did not correlate with the increase in personal space size during the pandemic (all *p* > .148).

Prior work has shown that human “intruders” within a subject’s personal space increase discomfort of the subject at progressively closer distances (4, 7, 12-14), perhaps following a power law function (8). To test whether such intrusion-driven discomfort levels changed during the pandemic, subjects were asked to rate their discomfort in response to real and virtual humans presented at various distances (25%, 50%, 100%, 200%, 400% of each subject’s personal space size), both before and during the pandemic. During the pandemic, discomfort to personal space intrusions increased significantly compared to the pre-pandemic discomfort ratings in response to both real and virtual humans, following a power law (Real: *p* < 0.0001, KS statistic 0.53; Virtual: *p* <0.0001; KS statistic 0.21; Fig. 2C, Supplementary Materials).

## Discussion

These results suggest that the COVID-19 pandemic led to a significant enlargement of the boundaries of personal space. Crucially, this personal space enlargement was evident even when there was no increase in infection risk, in a virtual reality environment. Moreover, discomfort ratings increased within the enlarged personal space. This pandemic-related increase in personal space may reflect a corresponding change in the neural representation of the portion of space that immediately surrounds the body -a “safety zone” requiring protection from harm (15). The magnitude of this enlargement was predicted by the *perceived* risk of being infected with COVID-19. Thus, beliefs about the infectiousness of the virus may have translated into an unconscious increase in the distance reflexively maintained from others, even in response to virtual humans.

Taken together, these findings suggest that the social distancing practiced during the COVID-19 pandemic altered the function of sensorimotor circuits of the brain that are involved in maintaining our physical safety. As the pandemic subsides, these mechanisms may revert to their prior set points. Alternatively, this change may persist for some unknown length of time, and influence how we interact in public and private settings. There is little historical precedent on which to base our expectations.

## Data Availability

All data are available upon request; please contact Daphne Holt at for data files.

## Acknowledgements

This work was supported by grants to DJH from the National Institute of Mental Health (5R01MH109562) and the Research Scholar Program of the Executive Committee on Research (ECOR) of Massachusetts General Hospital.

## References

1. L. A. Hayduk, Personal space: An evaluative and orienting overview. Psychological Bulletin 85, 117–134 (1978).

2. D. Uzzell, N. Horne, The influence of biological sex, sexuality and gender role on interpersonal distance. Br J Soc Psychol 45, 579–597 (2006).

3. H. Hecht, R. Welsch, J. Viehoff, M. R. Longo, The shape of personal space. Acta Psychol (Amst) 193, 113–122 (2019).

4. R. Welsch, C. von Castell, H. Hecht, The anisotropy of personal space. PLoS One 14, e0217587 (2019).

5. A. Sorokowska et al., Preferred Interpersonal Distances: A Global Comparison. Journal of Cross-Cultural Psychology 48, 577–592 (2017).

6. T. Iachini et al., Peripersonal and interpersonal space in virtual and real environments: Effects of gender and age. Journal of Environmental Psychology 45, 154–164 (2016).

7. L. A. Hayduk, The Permeability of Personal Space. Canadian Journal of Behavioural Science 13, 274–287 (1981).

8. R. B. H. Tootell et al., Psychological and Physiological Evidence for an Initial ‘Rough Sketch’ Calculation of Personal Space. Manuscript Under Review (2021).

9. L. A. Hayduk, Personal space: Where we now stand. Psychological Bulletin 94, 293–335 (1983).

10. M. Kaitz, Y. Bar-Haim, M. Lehrer, E. Grossman, Adult attachment style and interpersonal distance. Attach Hum Dev 6, 285–304 (2004).

11. L. Gerhold, COVID-19: Risk perception and Coping strategies. PsyArXi (2020).

12. J.S. LloberaB.,; Ruffini, G. ; Slater, M., Proxemics with multiple dynamic characters in an immersive virtual environment. ACM Transactions on Applied Perception 8, 1–12 (2010).

13. G. Schoretsanitis, A. Kutynia, K. Stegmayer, W. Strik, S. Walther, Keep at bay!-- Abnormal personal space regulation as marker of paranoia in schizophrenia. Eur Psychiatry 31, 1–7 (2016).

14. N. J. Felipe, R. Sommer, Invasions of personal space. Social Problems 14, 206– 214 (1966).

15. M. S. A. Graziano, D. F. Cooke, Parieto-frontal interactions, personal space, and defensive behavior. Neuropsychologia 44, 2621–2635 (2006).

