## Supplemental Materials for "Personal space Increases during the COVID-19 Pandemic in Response to Real and Virtual Humans"

This file includes:

Extended Methods

a. Subjects
b. COVID-19 Safety Procedures
c. Immersive Virtual Reality System
d. Personal Space Measurement
e. Distance Manipulation
f. Measure of Pandemic-related Beliefs and Experiences
g. Statistical Analyses
h. Fitting the Power Law Functions

References

**Extended Methods**

a. **Subjects**

Nineteen healthy individuals (9 female, mean age = 30.6 ± 11.3 years) were recruited via online advertisement posted on the Massachusetts General Hospital (MGH) Rally Website and enrolled in the study before the onset of the COVID-19 pandemic in Boston, MA (September 2019 - early March 2020). Inclusion criteria included: 1) age 18-55 years, 2) having normal or corrected to normal vision, and 3) having no unstable medical illness, active substance abuse or psychiatric disorder in the past 6 months (as assessed by (1)). Subjects were screened for virtual reality (VR) sickness using the Simulator Sickness Questionnaire (2), after spending approximately 10 minutes in the immersive VR system. No subjects were excluded due to VR sickness. Written informed consent was obtained from all subjects prior to enrollment, in accordance with the guidelines of the Partners Healthcare Institutional Review Board.

Twelve of the subjects described above (5 female, mean age = 33.3 ± 11.2 years) returned to complete a second visit, that was virtually identical to the first, during the COVID-19 pandemic (July 2020 - December 2020). All subjects who were able to return were enrolled in the second session. Subjects completed all of the same procedures during the two independent visits (which were an average of 10.04 ± 1.6 months apart).

In addition, a previously assessed sample of nine healthy individuals (5 female, mean age 27.1 ± 5.3 years), who had been recruited and consented in the same manner as the above sample, participated in two independent sessions before the beginning of the COVID-19 pandemic (an average of 13.5 ± 1.9 months apart) during which measurements of personal space with respect to real (but not virtual) humans were collected.

b. **COVID-19 Safety Procedures during the Second Session**

For research visits occurring during the pandemic, subjects were screened for COVID-19 symptoms and travel within 48 hours of arrival in accordance with MGH guidelines. In addition, mask wearing and social distancing policies were in effect for all subjects and staff members throughout the visits. The only exception to this (approved by the MGH COVID safety team) was during the measurement of personal space to real humans, when the subject wore a mask and protective eye goggles while the “experimenter” (a staff member, see below) did not wear a mask. This was done in order to maintain the same conditions from the perspective of the subject across the two visits (which occurred before and during the pandemic). Immediately after this procedure, the staff member/experimenter resumed wearing a mask.

c. **Immersive Virtual Reality System**

The HTC VIVE Virtual Reality System was used for the virtual reality (VR) portions of the study. The head-mounted display (HMD) presented stereoscopic images at a resolution of 1080 x 1200 pixels per eye, with a 110° field of view at a refresh rate of 90Hz. A software program designed to measure personal space (developed by Productive Edge <https://www.productiveedge.com/> ) was run via a SteamVR platform on an Alienware 15 R3 Laptop. In the HMD, the program displayed a neutral room (see Figure 1) in which virtual humans could be placed at different distances from the subjects and could appear to walk towards subjects while maintaining eye contact with them.

d. **Personal Space Measurement**

Personal space size was measured with respect to both real and virtual humans using the well-validated Stop Distance Procedure (SDP), during “passive” and “active” trials.

The SDP procedures using humans:

*Passive trials:* In the passive trials (which were not intermixed with the active trials), the subjects were asked to stand still while facing a real human (i.e., a staff member, the “experimenter”), standing 3 meters away from them. Subjects were instructed to maintain eye contact with the experimenter, who maintained a neutral facial expression, and were told that the experimenter would start walking slowly towards them, and that they should say “okay” when the experimenter reached the distance that the subject would normally maintain from such a person who they had just met (conversational distance; D1, passive). This distance was then measured. For these trials, the experimenters were trained to walk at approximately 0.1m/s.

*Active trials:* The active trials of the task began similarly, with the subjects standing 3 meters away from the human experimenter. However, in the active version of the procedure, the subjects were instructed to approach the experimenter, and to stop at the conversational distance described above and say “okay”. Again, the subjects were asked to maintain eye contact with the experimenter, who maintained a neutral facial expression. This distance was measured and recorded (D1, active).

Both the active and passive trials were conducted with a male and a female experimenter in a counterbalanced order.

The SDP procedures using avatars:

The same procedures were conducted in an immersive VR environment, using four different avatars (two males and two females). In the VR environment, the height of each avatar was set to equal the height of the subject, and the approach speed was set at 0.1m/s.

Each version of the SDP was repeated twice per stimulus in both modalities (real and virtual). Across subjects and time points, the following was counterbalanced: 1) the order of SDP modality (real and virtual), 2) the presentation order of the stimuli (two humans and four avatars), and 3) the passive and active SDP trial block.

e. **Distance Manipulation**

For each individual subject, personal space size was calculated separately for each modality (real and virtual) based on the average D1 of the active trials of that visit (since SDP measurements of active trials are slightly more stable than passive ones (3)). Multiples of each individual subject’s personal space size were calculated: 25%, 50%, 100%, 200%, 400%.

In two separate procedures, real and virtual humans were presented to the subject at these 5 different distances, in a counterbalanced, pseudorandomized order. For each trial, the subject began with their eyes closed, and then was asked to open their eyes during the presentation of each stimulus.The subject was instructed to stand still during the stimulus presentation and maintain eye contact with the real or virtual human. During each presentation, subjects were asked to rate their agreement to the statement “I want to move away” (discomfort ratings) on a Likert scale from 1-5 (1: not at all, 3: somewhat, 5: very much). The order of modality was the same as the order used for the personal space measurement within each subject and visit (3).

f. **Measure of Pandemic-related Beliefs and Experiences**

During the second visit, which occurred during the COVID-19 pandemic, subjects completed a validated self-report questionnaire that assessed their beliefs and feelings about the pandemic: the COVID 19: Risk Perception and Coping Strategies scale (4). This scale assessed subjects’ perceived risk of being infected with COVID-19 and anxiety and depressive symptoms related to the pandemic. These items were rated on a 5-point Likert scale.

g. **Statistical Analyses**

Repeated measures ANOVAs (modality X time) and paired samples t-tests were used to test for differences in personal space size and discomfort ratings across modality (real and virtual) and the two time points. Significance values of paired t-tests comparing discomfort ratings across distance percentages were corrected for multiple comparisons (Bonferroni, a = .05, corrected), within each time point and modality. Change scores were calculated as the difference between values collected at the second and first time point (POST minus PRE values, i.e., “after” minus “before” the onset of the COVID-19 pandemic). Thus a positive change score indicated an increase in the respective measure over time. Pearson’s correlations were used to measure relationships between changes in personal space size over time and beliefs and experiences related to the pandemic.

h. **Fitting the Power Law Functions**

Power law functions as $D=a x^{b}$ were fitted to the pooled discomfort ratings, where $D$ is the reported discomfort level, $x$ is the distance between the subject and the real or virtual human (as percentage of pre-pandemic personal space size), and $a, b$ (the prefactor, and the exponent, respectively) are parameters obtained by minimizing the sum squared error between the power law function and the data. Separate power law functions were fitted to pre-pandemic and pandemic data, for real and virtual humans (Figure 2D).

To test whether the power law functions were significantly different before versus during the pandemic, the fitting procedure was repeated 1000 times in each case, by bootstrapping the data with substitution. The resultant pre-pandemic and post-pandemic $a$ and $b$ parameters were compared using the nonparametric two-sample Kolmogorov-Smirnov (KS) test, separately for real and virtual humans. The prefactor $a$ was significantly different between the pre- and post pandemic timepoints, for both real and virtual humans (*p* < 0.0001 for both, KS statistics 0.36 and 0.20, respectively). Likewise, the exponent $b$ was significantly different between the pre- and post-pandemic timepoints for both real and virtual humans (*p* < 0.0001 for both, KS statistics 0.53 and 0.21, respectively).

**References**

1.	D. V. L. Sheehan, Y.; Harnett Sheehan, K.; Janavs, J.; Weiller, E.; Keskiner, A.; Schinka, J.; Knapp, E.; Sheehan, M.F.; Dunbar, G.C., The validity of the Mini International Neuropsychiatric Interview (MINI) according to the SCID-P and its reliability. *European Psychiatry* **12**, 232-241 (1997).

2.	S. Bruck, P. Watters, Cybersickness and Anxiety During Simulated Motion: Implications for VRET. *Stud Health Technol Inform* **144**, 169-173 (2009).

3.	R. B. H. Tootell *et al.*, Psychological and Physiological Evidence for an Initial ‘Rough Sketch’ Calculation of Personal Space. *Manuscript Under Review* (2021).

4.	L. Gerhold, COVID-19: Risk perception and Coping strategies. *PsyArXi* (2020).
